# The relation between macro- and microscopic intrinsic muscular alterations of the medial gastrocnemius in children with spastic cerebral palsy

**DOI:** 10.1101/2024.04.05.24305325

**Authors:** C. Lambrechts, J. Deschrevel, K. Maes, A. Andries, N. De Beukelaer, B. Hanssen, I. Vandekerckhove, A. Van Campenhout, G. Gayan-Ramirez, K. Desloovere

## Abstract

Children with spastic cerebral palsy (CP) are characterized by altered muscle growth, secondary to the pathological neural input to the muscular system, caused by the primary brain lesion. As a result, their medial gastrocnemius is commonly affected and is characterized by altered macro and microscopic muscular alterations. At the macroscopic level, the muscle volume (MV), anatomical cross-sectional area of the muscle belly (Belly-CSA), muscle belly length (ML), and the intrinsic muscle quality are reduced. At the microscopic level, the cross-sectional area of the fiber (Fiber-CSA) is characterized by an increased within-patient variability (coefficient of variation (CV)), the fiber type proportion is altered and capillarization is reduced. However, the relations between the muscular alterations at the macro- and microscopic level are not yet known. Therefore, this cross-sectional study integrated macro- and microscopic parameters of the medial gastrocnemius in one cohort of young ambulant children with CP and age-matched TD children, and explored how deficits in macroscopic muscle size are associated with alterations at the microscopic level. A group of 46 children with CP (median age 5.4 (3.3) years) and a control group of 34 TD children (median age 6.3 (3.4) years), who had data on microscopic muscular properties (defined through the histological analyses of muscle biopsies), as well as macroscopic muscle properties (defined by 3D freehand ultrasound) were included. We defined Pearson or Spearman correlations, depending on the data distribution. The macroscopic muscle size parameters (MV, Belly-CSA, ML) showed significant moderate correlations (0.504-0.592) with the microscopic average Fiber-CSA in TD and CP. To eliminate the common effect of anthropometric growth at the macro- as well as microscopic level, the data were expressed as deficits (i.e. z-scores from normative centile curves or means) or were normalized to body size parameters. A significant but low correlation was found between the z-scores of MV with the z-scores of the Fiber-CSA (r=0.420, p=0.006). The normalized muscle parameters also showed only low correlations between the macro- and microscopic muscle size parameters, namely between Belly-CSA and Fiber-CSA, both in the TD (r=0.408, p=0.023) and the CP (ρ=0.329, p=0.041) group. Explorations between macroscopic muscle parameters and other microscopic muscle parameters (capillary density, capillary to fiber ratio, and fiber type proportion) revealed no or only low correlations. These results highlighted the complexity of the interacting network of intrinsic muscle structures, with mainly low associations between the macro- and microstructural level, and it remains unclear how alterations in microscopic muscle structures contribute to the macroscopic muscle size deficits in children with CP.

## 1 INTRODUCTION

Skeletal muscles are the executive organs for locomotion and are therefore of indispensable value in daily live, as they allow us to walk, jump, run etc. However, in neurological disorders, such as cerebral palsy (CP), skeletal muscle function is commonly affected as a result of a brain lesion. CP describes a series of non-progressive disorders caused by an injury that occurs in the developing fetal or infant brain, up to 2 years after birth (Rosenbaum et al., 2007). With a global prevalence of 1.6 to 3.4 per 1000 live births, CP is the most common cause of motor impairment in children (McIntyre et al., 2022), with spastic CP being the predominant subtype, accounting for approximately 80% of the CP population (Michael-Asalu et al., 2019). Based on their functional abilities, children with CP are assigned to one of the five levels of the gross motor function classification system (GMFCS), with a higher level reflecting a more severe functional impairment (Palisano et al., 1997), and level I, II and III representing children who are ambulant. Spasticity, muscle weakness, muscle stiffness and limited joint range of motion are frequently described neuromuscular symptoms, some of which may increase over time. The pathological neural drive from the brain to the muscle is the primary contributor to these neuromuscular symptoms, causing secondary progressive macroscopic and microscopic muscular changes, given the high plasticity of skeletal muscle tissue (Graham et al., 2016; Handsfield et al., 2022).

In recent years, macroscopic muscle morphology has been increasingly studied, supported by improved accessible muscle imaging techniques, such as the three-dimensional freehand ultrasound (3DfUS), which enables valid and reliable descriptions of the muscle structure (Hanssen et al., 2023; Cenni et al., 2018). The gastrocnemius muscle has been most often investigated (Williams et al., 2021), probably because of its major and relevant contribution to daily functional activities and its susceptibility to spasticity and weakness in children with CP (Neptune et al., 2001; Mathewson and Lieber, 2015). We recently described the longitudinal trajectory of morphological alterations in the medial gastrocnemius throughout growth in ambulant children with CP, aged between 6 months and 11 years (De Beukelaer et al., 2023). To account for muscular changes caused by anthropometric growth, muscle volume (MV) and cross-sectional area of the muscle belly (Belly-CSA) were normalized to mass and/or height. Normalized MV (nMV) and Belly-CSA (nBelly-CSA) showed reduced growth rates compared to age-matched typically developing (TD) peers, and these reductions depended on the severity of the child’s motor impairment. Also other investigations, based on cross-sectional designs, have identified nMV deficits of 24-52%, and normalized muscle belly length (ML) deficits, with ranges of 9-19% in children with CP aged between 2 to 12 years compared to age-matched TD children (Malaiya et al., 2007; Barber et al., 2011, 2016; Obst et al., 2017; Schless et al., 2018, 2019; Hanssen et al., 2021). Moreover, an indirect sign of reduced intrinsic muscle quality, expressed by increased ultrasound echo-intensity (EI) (i.e. the averaged gray-scale value of ultrasound images), has been observed in the CP compared to the TD population (Obst et al., 2017; Schless et al., 2018, 2019). It can thus be concluded that there is ample evidence of hampered muscle growth resulting in reduced muscle size in children with CP compared to TD children.

At the microscopic level, several muscular alterations have already been reported in CP but results were inconsistent between studies. This is related to the fact that these studies included a heterogeneous group of CP participants in terms of age and functional ability, and used inappropriate controls (not age- and muscle-matched) for comparison. Additionally, they were mainly conducted in adolescents or young adults with established muscle contractures (Smith et al., 2013; De Bruin et al., 2014). This is probably due to the difficulty of collecting muscle biopsies, especially in young children and more particularly in age-matched TD children. To partially encounter these methodological problems, we adapted an existing micro biopsy technique (Deschrevel et al., 2023b), and recently collected biopsies of the medial gastrocnemius (MG) in an extended group of children with CP and age-matched TD children. This allowed to comprehensively describe the microscopic MG properties of 46 ambulant children with CP, aged 2 to 9 years, and from 45 age-matched TD children (Deschrevel et al., 2023a). We thereby found that the muscle fibers, which are the multinucleated cells of the skeletal muscle, were characterized by a lower percentage of type I fibers (65% TD vs 52% CP) and a higher amount of type IIx fibers (7% TD vs 22% CP) in children with CP compared to their TD peers (Deschrevel et al., 2023a). This was in contrast to the type I fiber predominance that was previously reported for the MG of children and adolescents with CP (Ito et al., 1996; Rose et al., 1994), but this conclusion was based on the comparison with another muscle type of TD children (Rose et al., 1994) or with muscles from previous autopsy studies (Ito et al., 1996). We further found a higher coefficient of variation (CV, which expresses within-subject variability in fiber size) in the MG of children with CP (Deschrevel et al., 2023), which was also a consistent finding in several earlier studies performed on different muscles (Rose et al., 1994; Ito et al., 1996; Marbini et al., 2002; Kahn et al., 2023). More specifically, we found that children with CP had a significant higher percentage of smaller fibers (i.e. a fiber area between 0-750 µm2) compared to TD children (30±26% in CP vs 8±18% in TD) while the averaged cross-sectional area of all fibers, expressed as absolute values (Fiber-CSA) or as normalized values (i.e. scaled to squared fibula length to account for anthropometric growth) (nFiber-CSA) was not altered (Deschrevel et al., 2023a). These findings were not entirely in line with previous findings, since some earlier studies showed a smaller averaged Fiber-CSA in children with CP compared with TD children (Borg et al., 2019; Kahn et al., 2023), while one study, which compared different muscle types for the CP and TD group, did not find any changes in children with CP (Rose et al., 1994). In the same population, we found a lower capillary to fiber ratio (C/F) and a reduced capillary fiber density in the MG of children with CP compared to TD children (Deschrevel et al., 2023a). This was in agreement with the 38% lower capillary density reported in the biceps brachii muscle in young adults with CP compared to TD controls (Pontén and Stål, 2007). Taking together, these data indicated that several microscopic alterations are clearly present in the MG of children with CP, but vary a lot between subjects and between studies. The inconsistencies are most-likely related to the often inappropriate control groups used for comparison.

So far, macro- and microscopic muscular properties have not yet been investigated in combination. Hence, the interaction between alterations at the macro- and microscopic level remains unknown. A better knowledge on this interaction may potentially help to better understand the observed clinical symptoms in children with CP (Mathewson and Lieber, 2015; Multani et al., 2019; Howard and Herzog, 2021). In particular, macroscopic muscle growth deficits were found to be associated to clinical symptoms such as spasticity (Peeters et al., 2023), muscle weakness (Hanssen et al., 2021) and the development of contractures (Gough and Shortland, 2012; Pingel et al., 2017; Handsfield et al., 2022), leading to functional problems such as deteriorated gait (Schless et al., 2019). However, the potential role of microscopic muscle alterations within this complex interacting network of muscular alterations and clinical symptoms remains unknown. This requires a combined dataset of macro- and microscopic muscle properties defined from one patient cohort. Our research group recently established such a unique combined database, since all children with CP and TD children who were enrolled in the above described study on microscopic properties where MG biopsies were collected (Deschrevel et al., 2023a), also received 3DfUS of the MG to assess macroscopic properties. This created the opportunity to unravel the interaction between macro- and microscopic intrinsic muscle characteristics.

Therefore, the overall aim of the current study was to investigate whether intrinsic macroscopic muscle properties of the MG are correlated with muscle properties at the microscopic level. To achieve this goal, we examined a unique combined MG muscle database, covering both the macro- and microscopic properties in 2 to 9 years old children with CP and age-matched TD children. Within this correlation study, we transformed the outcomes to exclude the impact of anthropometric growth on the correlations between macro- and microscopic properties. First, we quantified associations between the macroscopic muscle size parameters (muscle volume, CSA of muscle belly, muscle belly length) with microscopic muscle size parameters (CSA of muscle fibers and its CV). As a primary hypothesis, we stated that alterations in the macroscopic muscle size would be significantly associated with alterations in the microscopic muscle fiber size in children with CP. We also explored whether the associations between macro- and microscopic muscle size parameters differed between children with CP and TD children. Finally, we also explored potential associations between other macro- and microscopic properties (such as echo-intensity, fiber type proportion and capillarization), which may suggest novel underlying mechanisms to be further investigated in future research.

## 2 MATERIALS AND METHODS

### 2.1 Participants

This cross-sectional study is based on the same study population as reported by Deschrevel et al., 2023a, which included children with CP and TD children who had a biopsy collection. For the current study, only children who had a successful 3DfUS measure, next to their biopsy collection, were included. The CP group from the original study included 46 ambulant children diagnosed with spastic CP, aged between 2 to 9 years, with a GMFCS classification level I, II or III, and with uni- or bilateral involvement. The children were recruited through the Cerebral Palsy Reference Centre and the Clinical Motion Analysis Laboratory of the University Hospitals Leuven (Belgium). The following exclusion criteria were applied: (1) diagnosis of dystonia or ataxia, (2) previous botulinum toxin type A (BoNT-A) injections in the past 6 months prior to the study assessments, (3) history of orthopaedic surgery during the past 2 years, (4) previous surgery on the gastrocnemius muscle at any time point, (5) severe co-morbidities, such as cognitive problems, that prevent accurate assessments. All these children with CP had a successful 3DfUS measure, apart from the muscle biopsy and could thus be enrolled in the current study. The TD group from the original study included 45 age-matched children who had a muscle biopsy, of which 34 children had also a successful 3DfUS measure and could thus be enrolled in the current study. These TD children were recruited through the University Hospitals Leuven (Belgium) if they were planned for surgery with anaesthesia such as removal of orthopaedic implants of the upper limbs, or for ear-nose-throat surgery. The exclusion criteria were (1) requirement of lower limb surgery, (2) a history of musculoskeletal or neurological disorders, and (3) engaging in sport activities more than 3 hours per week. Written informed consent was given for all children from their parents or legal guardians. Ethical approval was obtained from the Ethical Committee of the University Hospitals of Leuven (S61110 and S62645).

For all enrolled children, macroscopic properties were evaluated with 3DfUS and after biopsy collection, microscopic properties were determined on histological staining.

### 2.2 Muscle biopsy collection to define microscopic properties

Muscle biopsy collection and analysis were performed, as described previously (Deschrevel et al., 2023b). Briefly, percutaneous microscopic biopsies of the MG were collected via guided ultrasonography during procedures that required general anaesthesia (as specified above). For the ultrasonography guided biopsy, a disposable core biopsy instrument (Bard® Mission™ Disposable Core Biopsy Instrument 16G) was used and inserted in the mid-belly of the MG, as illustrated in Figure 1A. The biopsies were collected from the most affected leg (defined by the clinical scores for MG spasticity and weakness) for children with CP, while for TD children, a random leg was selected. All biopsies were taken by experienced paediatric orthopaedic surgeons. MG biopsies were cut into five μm slices by using a cryostat, and transferred to a charged glass slide (SuperFrost, VWR). Thereafter, the slices were stored at -20° until further use. For histological analyses, slides were stained with hematoxylin and eosin to qualitatively evaluate the cross-sectional orientation of the sections. Next, myosin heavy chain (MHC) immunostaining was used to distinguish between the different fiber types to determine the fiber type proportion and average fiber CSA (Fiber-CSA). The coefficient of variation (CV), which expresses fiber size variability per subject, was calculated by dividing the standard deviation (SD) of the Fiber-CSA by the average values of the Fiber-CSA, multiplied by 100 (CV=(SD/mean)*100). Finally, capillary density (number of capillaries/mm²) and C/F (amount of capillaries surrounding one fiber) were determined by using antibodies against capillaries (CD31) and type I fibers.

**Figure 1.**
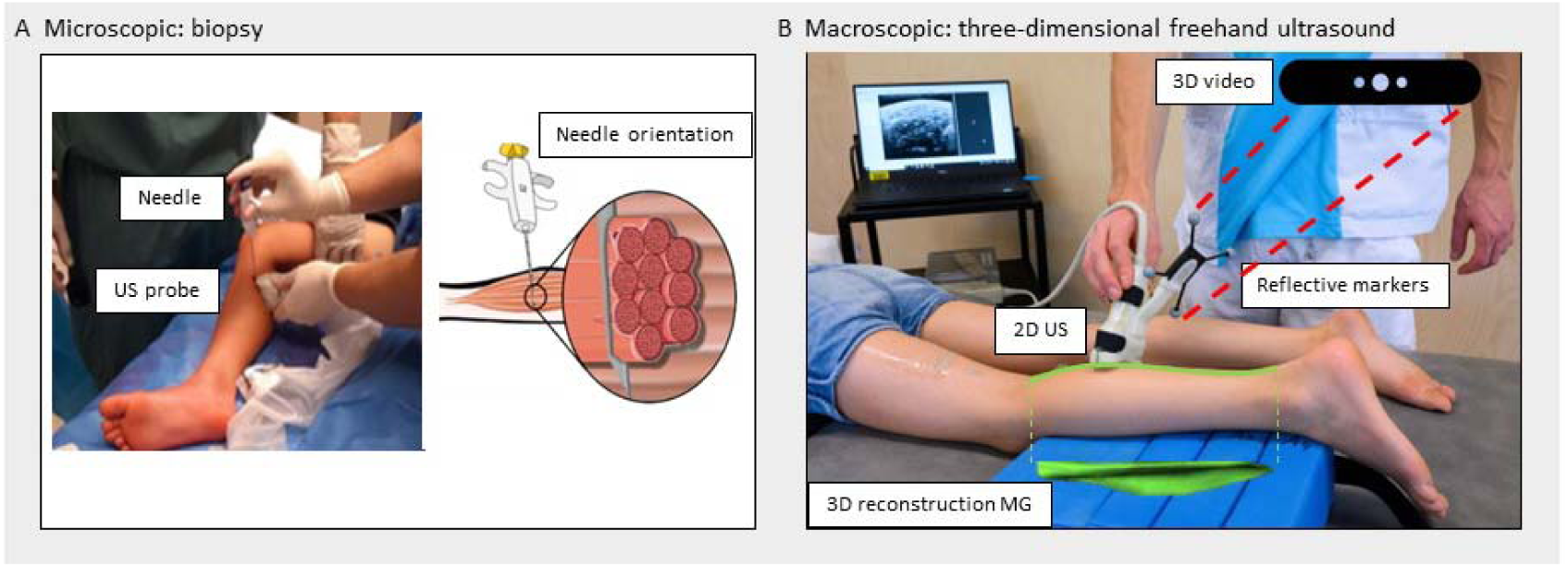
A) Overview of ultrasound (US) guided biopsy collection B) Overview of three-dimensional freehand ultrasound (3DfUS) measurement set-up

### 2.3 Three-Dimensional Freehand Ultrasonography to define macroscopic properties

3DfUS was applied to construct 3D images of the MG, as described earlier (Hanssen et al., 2023). Briefly, 3DfUS combines two-dimensional B-mode ultrasonography (TelemedEchoblaster 128 Ext-1Z, with a 5-9 cm 10-MHz linear US transducer, Telemed Ltd., Vilnius Lithuania) with motion tracking of the ultrasound probe (Optitrack V120:Trio, NaturalPoint Inc., Corvallis, Oregon, USA). The motion of four markers attached to the ultrasound probe were tracked while sweeping over the MG muscle belly, to define the position and orientation of each two-dimensional ultrasound image. These data were used to create a 3D reconstruction of the MG muscle. The 3DfUS were performed on the same leg that was selected for biopsy collection. The participants lay in prone position on a table with their lower leg supported and foot in a neutral resting position, not touching the table, as illustrated in Figure 1B. Data was extracted and processed with STRADWIN software (version 6.0; Mechanical Engineering, Cambridge University, Cambridge, UK). By indicating landmarks (origin, muscle-tendon junction, insertion) and manually marking transverse segmentations along the inner muscle border, macroscopic muscle parameters were estimated. We defined MV (ml) as outcome for the 3D size of the MG muscle belly, the anatomical Belly-CSA (mm^2^) as the 2D cross-section of the mid-belly perpendicular to the longitudinal axis of the muscle, and the ML (mm) as the distance between the most superficial part of the medial femur condyle and the muscle-tendon junction. To estimate the intrinsic muscle quality, the averaged EI-value of the whole 3D reconstruction was calculated. The 3DfUS measurements and processing were performed by well-trained trained researchers with more than 4 years of experience.

#### Primary and secondary outcomes

Because muscle size parameters increase with anthropometric growth, some outcomes needed to be normalized. Normalization was performed following previously reported approaches for ratio scaling (De Beukelaer et al., 2023; Deschrevel et al., 2023a). We normalized the MV to the product of body mass and height (nMV, ml/kg*m), the Belly-CSA and Fiber-CSA to the squared fibula length (nBelly-CSA, mm^2^/fibula^2^ and nFiber-CSA, µm^2^/fibula^2^), and ML to the height of the child (nML, mm/m). However, we recently found that these normalization techniques work well for certain age ranges, but are imperfect for wide age ranges (De Beukelaer et al., 2023; Vandekerckhove et al., 2023). Moreover, visual inspection of all TD data from the current study with respect to anthropometric parameters highlighted that also other outcomes (such as the capillary density and C/F) altered with the natural growth of the child. As an alternative for normalization, z-scores can be used. We recently introduced the use of anthropometric-related percentile curves of TD children to convert muscle size outcomes of the children with CP into z-scores, which reflect alterations as deficits that are independent from anthropometric growth (Vandekerckhove et al., 2023). These non-dimensional z-scores were found to successfully describe progress in macroscopic muscle alterations during growth in children with CP. Therefore, in the current study, besides the use of normalized outcomes (as commonly reported in previous studies), we also defined z-scores for each muscle property for children with CP, using our established TD databases as a reference. For all muscle parameters that showed changes with anthropometric growth in TD children, z-scores were defined based on anthropometric-related percentile curves. These percentile curves were developed using generalized additive models for location, scale and shape (GAMLSS) in R (Rigby and Stasinopoulos, 2005; Borghi et al., 2006; Stasinopoulos and Rigby, 2007), as explained in more detail by Vandekerckhove et al., 2023. Z-scores of macroscopic muscle size parameters (zMV, zBelly-CSA, zML) were calculated using percentile curves that were based on 154 TD children from our established muscle database (Vandekerckhove et al., 2023). Z-scores of microscopic parameters that showed changes with anthropometric growth in TD children (zFiber-CSA, zCapillary density, zC/F) were calculated using percentile curves that were based on 34 TD children (i.e. our current TD microscopic muscle database). Finally, the CV of the Fiber-CSA (which remained stable in growing CP children) was also presented as z-scores (zCV) by subtracting the mean of the current TD sample, divided by the SD of the 34 TD children.

The <u>primary outcomes</u>, to test the study hypothesis, were the z-scores for five main muscle size parameters, namely zMV, zBelly-CSA and zML for the macroscopic parameters, and zFiber-CSA and zCV, for the microscopic parameters. However, to facilitate comparison to previous studies and to also explore the relation between macro-and microscopic muscle properties in TD children, we additionally investigated the absolute and normalized muscle size parameters as <u>secondary outcomes</u> (MV, Belly-CSA, ML, Fiber-CSA, CV, nMV, nBelly-CSA, nML, nFiber-CSA). Furthermore, next to the above mentioned muscle size parameters, we also investigated other macro- and microscopic parameters (EI, Capillary density, C/F, zCapillary density, and zC/F) as extra secondary outcomes.

### 2.4 Statistical analysis

For statistical analyses, normality was checked using the Shapiro-Wilk test and visual inspection of QQ plots. Normally distributed data are represented as mean with SD, and non-normally distributed data as median with interquartile range (IQR). For all analyses, the significance level was set at 0.05 and Bonferroni correction was applied in case of multiple testing.

A preparatory step involved <u>descriptive statistics</u>. We described the subject demographics, anthropometric data and macro- and microscopic muscular properties for the TD and CP group, whereby the differences between groups were tested with an independent T-test (with check of equality of variances) or a Mann-Whitney U test (in case of non-normal data distribution). As the between-group differences of the macro and microscopic muscular properties were not the main focus of the current study, these results are only briefly reported and summarized in supplementary tables.

For the <u>correlation analyses</u>, we used Pearson correlation coefficient (r) for normally distributed data, or the Spearman’s rho correlation coefficient (ρ) for not-normally distributed data. Correlation coefficients were interpreted as: <0.300 neglectable, 0.300-0.499 low, 0.500-0.699 moderate, 0.700-0.899 high and ≥0.900 very high (Dennis E. Hinkle, 2003). To test the study hypothesis (i.e. the association between the alterations in the macro- and microscopic muscle size parameters), the correlations of the z-scores of the three macroscopic muscle size parameters with the z-scores of the two microscopic muscle size parameters (i.e. the primary outcomes) were defined, with post-hoc Bonferroni correction for multiple testing (alpha 0.05/6=alpha 0.00833). For the correlations between secondary macro- and microscopic outcomes, correction for multiple testing was not applied, because of the exploratory nature of these additional analyses. Statistics were performed using IBM SPSS Statistics (version 29.0.2.0, Armonk, NY, USA). Graphs were made using Graphpad Prism (version 10.1.2, San Diego, CA, USA).

## 3 RESULTS

### 3.1 Subject demographics, anthropometric data and macro- and microscopic properties

The <u>study demographics and anthropometrics</u> are represented in Table 1 as median (IQR), since most parameters were not-normally distributed. All 46 children with CP who participated to the earlier biopsy study of (Deschrevel et al., 2023a), also had a successful 3DfUS measurement of MG and where therefore included for the current study. They were aged between 2.5 and 9.3 years (median (IQR) age of 5.4 years (3.3)). The CP group included 25 boys and 21 girls, 12 children were classified as GMFCS level I, 21 as GMFCS level II and 13 as GMFCS level III, and 14 children were diagnosed with unilateral and 32 with bilateral involvement. The sample included 24 BoNT-A naïve children and 22 children with a BoNT-A history at the moment of the biopsy collection and 3DfUS measurements. All children received regular physiotherapy on a weekly base, as well as orthotics, as part of their routine therapy. The final TD sample for the current study included 34 children who had a successful 3DfUS measurement next to the biopsy collection. The TD children were aged between 2.8 and 9.8 years (median (IQR) age of 6.3 years (3.4)) and there were 19 boys and 15 girls. The between-group analysis of the anthropometric characteristics revealed a significant younger age (-14%, p=0.038), lower body weight (-16%, p=0.007), body length (-11%, p<0.001), and fibula length (-9%, p=0.004) for children with CP compared to the TD children.

**TABLE 1.** Participant demographics and anthropometric data.

|  | TD | CP | p-MWU |
| --- | --- | --- | --- |
| N | 34 | 46 |  |
| Sex (boys/girls) | 19/15 | 25/21 |  |
| Age (years) | 6.3 (3.4) | 5.4 (3.3) | **0.038** |
| Age range (years) | 2.8-9.8 | 2.5-9.3 |  |
| Body weight (kg) | 21 (7.9) | 17.6 (8.3) | **0.007** |
| Body length (cm) | 122 (26) | 108.3 (23.9) | **<0.001** |
| Fibula length (cm) | 25.3 (5.7) | 23 (6.3) | **0.004** |
| GMFCS level (I/II/III) | NA | 12/21/13 |  |
| Involvement (uni/bilateral) | NA | 14/32 |  |
| BoNT naïve/history | NA | 24/22 |  |

All anthropometric parameters are displayed as median (interquartile range)
Abbreviations: TD, typically developing; CP, cerebral palsy; p-MWU, p-value of the Mann-Whitney U; kg, kilogram; cm, centimetre; GMFCS, Gross motor Function Classification System; BoNT, botulinum toxin type A
The critical p-value for differences between TD and CP is 0.05

For the <u>macroscopic muscle outcomes</u>, the means and SD’s for both groups, along with the results of the independent t-test between groups, are presented in Table S1 (supplementary material) while individual datapoints of all macroscopic parameters are illustrated in Figure S1 (supplementary material). All absolute and normalized macroscopic parameters showed significant differences between the CP and TD group (p<0.001). Children with CP, in comparison to TD children, had a reduced absolute (-44%) and normalized (-28%) MV, as well as a reduced absolute (-34%) and normalized (-19%) Belly-CSA. Similarly, deficits of 19% and 11% were found for the absolute and normalized ML, respectively, while EI was 13% increased. For <u>the microscopic muscle outcomes</u>, the medians and IQR of the TD children and children with CP, along with the results of the Mann-Whitney U test, are presented in Table S2 (supplementary material). Individual datapoints of all microscopic parameters are illustrated in Figure S2 (Supplementary material). The absolute Fiber-CSA tended to be lower in children with CP compared to TD children, however, this tendency was statistically not significant (after Bonferroni correction) and the nFiber-CSA was not significantly different between groups. The CV was significantly larger (p<0.001) in children with CP (+42% CV) compared to TD children. A fiber shift with less type I (-23%, p<0.001) and more type IIx (+307%, p<0.001) fibers was seen in children with CP compared to TD children. The amount of capillaries per squared millimetre and the C/F were significantly lower (-26%, p<0.001 and -46%, p<0.001, respectively) in children with compared to TD children.

### 3.2 Correlations between macro- and microscopic muscle parameters in TD and CP children

#### 3.2.a Correlations between macro- and microscopic muscle size

The correlations between the macro- and microscopic muscle size parameters, expressed as z-scores (primary outcomes) and as absolute values as well as normalized values (secondary outcomes), with corresponding p-values are listed in Table 2.

**TABLE 2.**
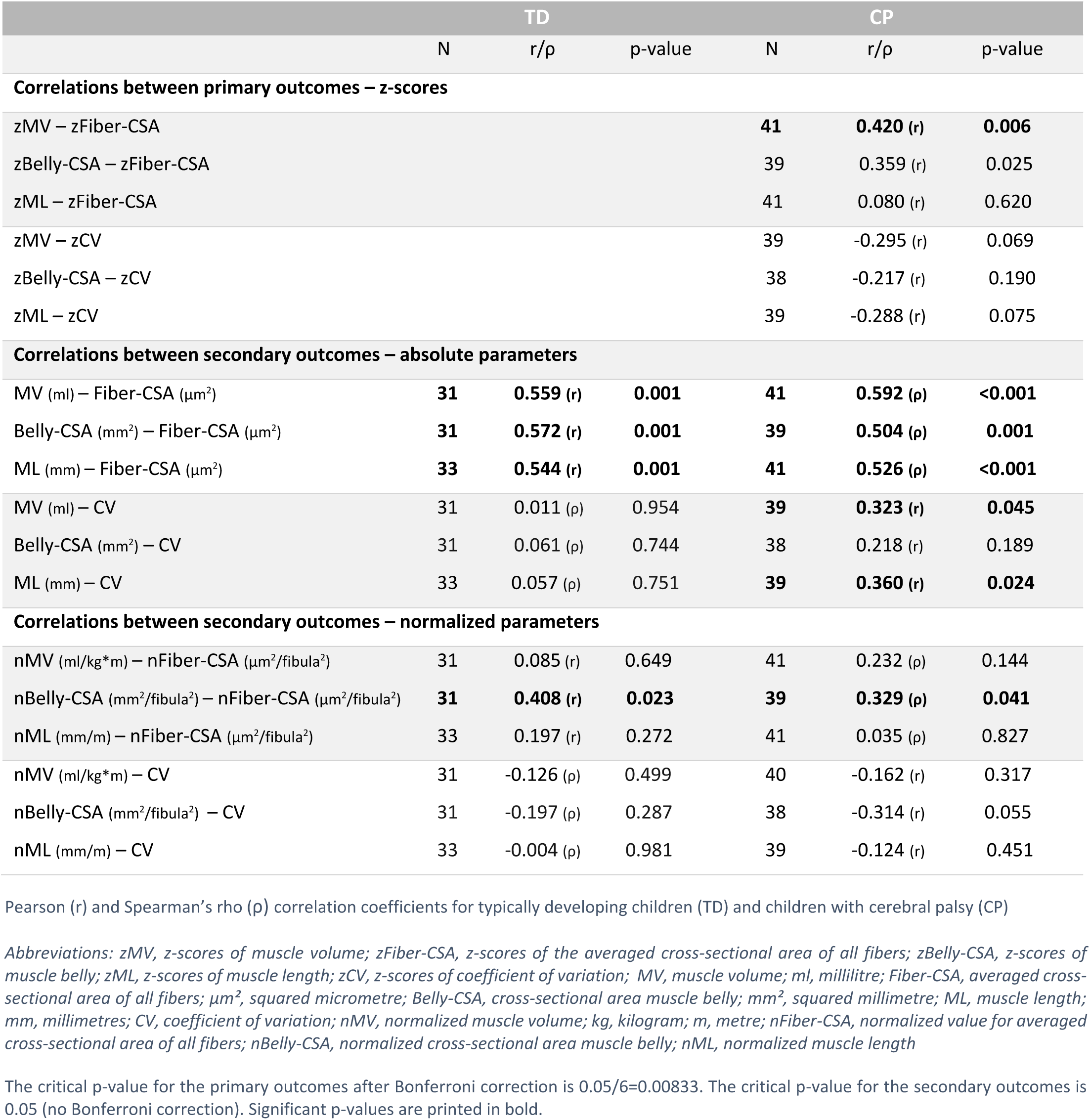
Correlations between macroscopic and microscopic muscle size parameters.

<u>For the primary outcomes,</u> the macroscopic zMV showed a significant but low correlation with the microscopic zFiber-CSA (r=0.420, p=0.006). The correlation between the macroscopic zBelly-CSA and the microscopic zFiber-CSA was significant (r=0.359, p=0.025), but was classified as low and not statistically significant after Bonferroni correction. The macroscopic zML showed no significant correlation with the microscopic zFiber-CSA. The correlations between all macroscopic outcomes (zMV, zBelly-CSA and zML) and the microscopic zCV were negligible.

<u>For the secondary outcomes</u>, moderate significant correlations were found between the macroscopic MV and microscopic Fiber-CSA in the TD (r=0.559, p=0.001) and CP (ρ=0.592, p<0.001) group. However, when eliminating the effect of anthropometric growth through normalization, these correlations were non-significant and negligible or low between nMV and nFiber-CSA in both groups (TD r=0.085, p=0.649; CP ρ=0.232, p=0.144). The macroscopic Belly-CSA was moderately correlated with the microscopic Fiber-CSA in the TD as well as the CP group (TD r=0.572, p=0.001; CP ρ=0.504, p=0.001). When normalizing those parameters, only low correlations were found between nBelly-CSA and nFiber-CSA in TD (r=0.408, p=0.023) as well as the CP group (ρ=0.329, p=0.041). The macroscopic ML showed significant moderate correlations with the microscopic Fiber-CSA (TD r=0.544, p=0.001; CP ρ=0.526, p<0.001), while negligible correlations were found between the normalized equivalents, i.e. between the macroscopic nML and the microscopic nFiber-CSA. Correlations between the absolute macroscopic MV and ML parameters and the microscopic CV parameter were low (r=0.323, p=0.045 and r=0.360, p=0.024, respectively) in the CP group, while all normalized macroscopic parameters (nMV, nBelly-CSA and nMV) showed low to negligible (non-significant) correlations with the microscopic CV. Scatterplots between the macro- and microscopic muscle size outcomes for the TD and CP group are presented in Figure 2 (except for the negligible correlations, i.e. when r/ρ <0.3).

**FIGURE 2.**
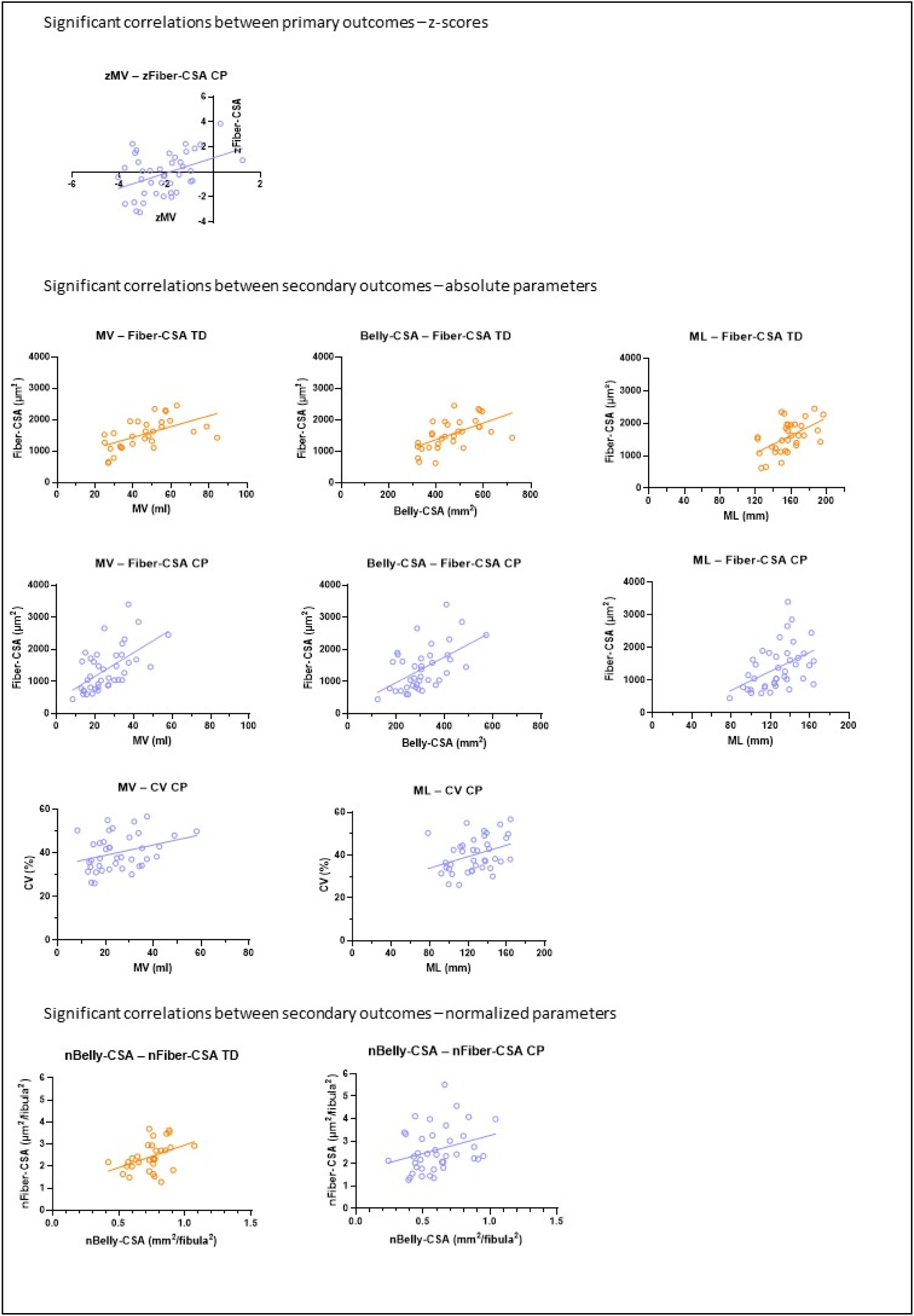
Scatterplots of individual datapoints for the significant correlations between the macro- and microscopic muscle size parameters in typically developing (TD) children (orange) and children with cerebral palsy (CP) (purple) Abbreviations: zMV, z-scores of muscle volume; zFiber-CSA, z-scores of the averaged cross-sectional area of all fibers; MV, muscle volume; Fiber-CSA, averaged cross-sectional area of all fibers; µm^2^, squared micrometre; Belly-CSA, cross-sectional area muscle belly; mm^2^, squared millimetre; ML, muscle length; mm, millimetres; CV, coefficient of variation; nBelly-CSA, normalized cross-sectional area muscle belly; nFiber-CSA, normalized value for averaged cross-sectional area of all fibers

#### 3.3.b Additional correlation analyses between macroscopic and microscopic muscle parameters

Here, we explored the correlations of the macroscopic size parameters with microscopic capillarisation parameters and fiber type proportion. We also explored the association of the macroscopic EI with microscopic parameters. The results of these secondary correlation analyses are presented in Table 3.

**TABLE 3.**
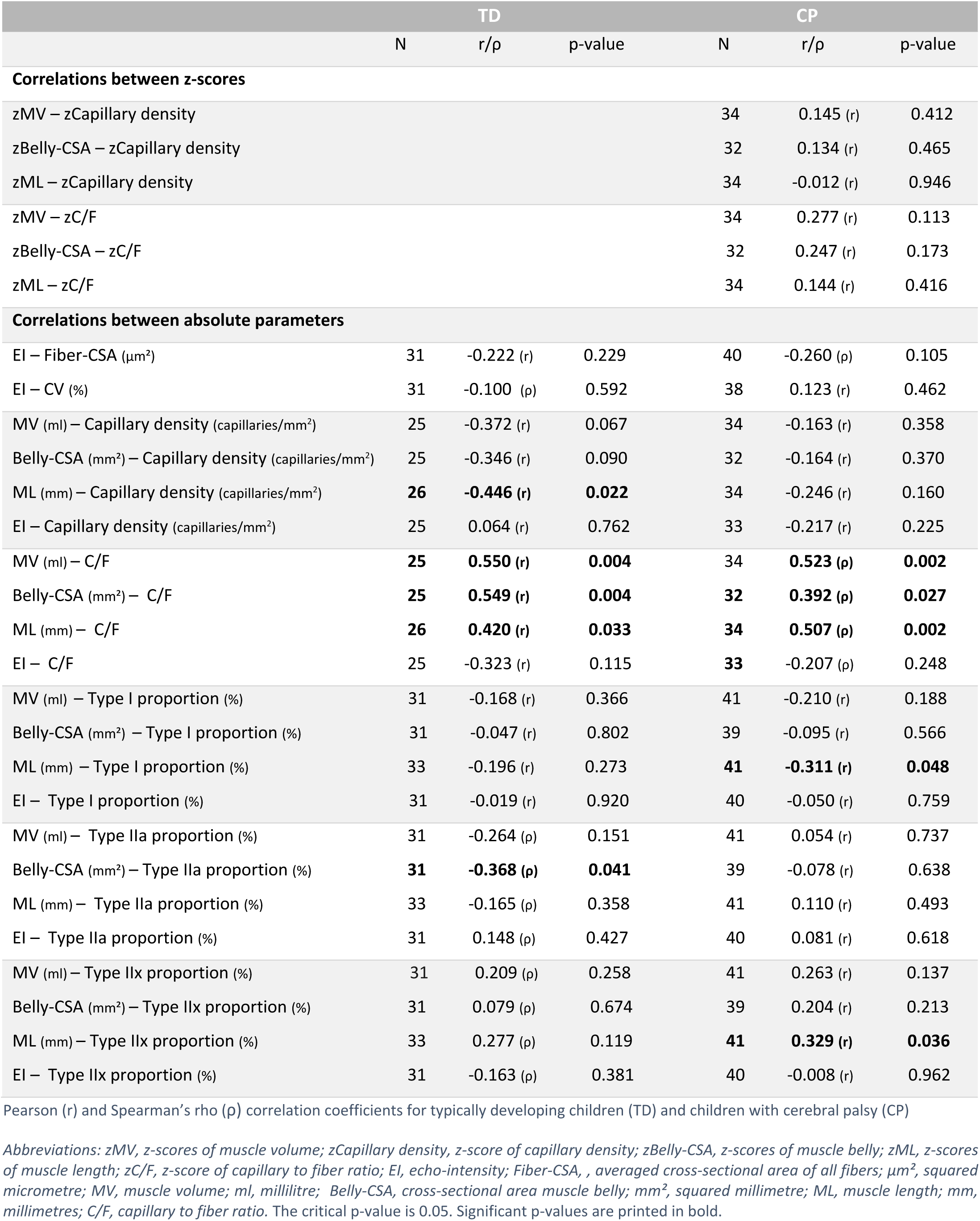
Additional correlation analyses between macroscopic and microscopic muscle parameters.

<u>For the z-scores</u>, there were no significant correlations between the macroscopic parameters and the microscopic capillarization (capillary density and C/F).

<u>For the absolute values</u>, we found significant moderate correlations between the macroscopic MV and the C/F for both groups (TD r=0.550, CP ρ=0.523). Significant correlations were also found between the macroscopic Belly-CSA and the C/F, which were moderate for TD children (r=0.549) and low for children with CP (ρ=0.392), as well as between the macroscopic ML and the C/F, which were low for TD children (r=0.420) and moderate for children with CP (ρ=0.507). Additionally, a significant but low correlation was also found between the macroscopic ML and the microscopic capillary density of the fibers (r=-0.446) for the TD children. Finally, absolute Belly-CSA was significantly correlated with fiber type proportion of type IIa (ρ=-0.368) in the TD group, and ML was significantly correlated with fiber type proportion of type I (r=-0.311) and IIx (r=0.329) in the CP group. Finally, EI showed non-significant negligible correlations with microscopic muscle properties (except for a low non-significant correlation with C/F (r=-0.323) in the TD group).

## 4. DISCUSSION

Overall, the results of the current correlation study on intrinsic muscular properties highlighted that the associations between altered macro- and microscopic properties of the MG in children with CP are only low to moderate.

The current study confirmed altered intrinsic muscle properties in children with CP compared to TD children. Indeed, at the macroscopic level, the investigated children with CP showed a reduced MG muscle size and increased values of EI (suggesting a lower intrinsic muscle quality), which is in line with previous literature on children with CP under 12 years of age (Malaiya et al., 2007; Pillen et al., 2007; Barber et al., 2011, 2016; Pitcher et al., 2015; Obst et al., 2017; Hanssen et al., 2021; De Beukelaer et al., 2023). At the microscopic level, the fiber CSA of the MG tended to be smaller, but especially showed variable sizes within patients, with reduced capillarization, and a shift to a faster fiber type, as described by Deschrevel et al. 2023a.

Although the alterations at the macro- and microscopic muscular levels were evident, their associations were limited. Hence, our hypothesis stating that macro- and microscopic muscle size parameters are related was only partially confirmed. The relation between the primary outcomes, namely the deficits in macro- and microscopic muscle size parameters with respect to TD control data (z-scores), indicated only one significant correlation, highlighting that increasing deficits in muscle volume (zMV) are weakly associated with increasing deficits in fiber CSA (zFiber-CSA). The results on the absolute values for muscle size revealed that the macroscopic muscle volume (MV), muscle CSA (Belly-CSA) and muscle length (ML) were moderately correlated to the microscopic Fiber-CSA, both in the TD and CP group. These correlations may be primarily attributed to the effect of anthropometric growth, since these correlations were not significant or much lower for the normalized and z-values. Indeed, growing children present with increasing muscle size at all structural levels, i.e. increasing MV, Belly-CSA and ML, as well as Fiber-CSA. These findings highlighted the importance of excluding the natural anthropometric growth of the muscle to investigate the clinical relevance of the relationship between growth dependent parameters. However, it should be noted that not all macroscopic morphological MG parameters have a linear relationship with anthropometric growth. Hence, the validity of normalization based on body size measurements (i.e. ratio scaling) may be questioned for groups with wide age-ranges (De Beukelaer et al., 2023; Peeters et al., 2023). Therefore, z-scores based on percentile growth curves of TD children were considered more valid to express the muscle deficits in children with CP with respect to TD data (Vandekerckhove et al., 2023). The use of z-scores resulted in slightly stronger correlations compared to the normalized parameters.

Overall, we expected higher correlation coefficients between the macro- and microscopic muscle size parameters. The current data suggest that muscle size deficits at the macroscopic level were not equally present at the microscopic level, highlighting the complexity of the interaction between muscular components. At the macroscopic level, the muscle volume is defined by the cross-sectional (Belly-CSA) and longitudinal size (ML) of the muscle, while at the microscopic level, we could only focus on the fiber cross-sectional size (Fiber-CSA). The muscle is built of contractile tissue (i.e. the muscle fibers), as well as the extracellular matrix. This extracellular matrix consists mainly of vasculature and connective tissue (collagen), and makes up to 20% of the entire muscle in healthy subjects (Haun et al., 2019). Contrary to our expectations, the strength of the correlations between the macro- and microscopic muscle sizes were more or less similar for the CP and the TD group. This was surprising, since children with CP, compared to TD children, tend to have more extracellular collagen content and fat infiltration (Noble et al., 2014; D’Souza et al., 2020; Smith et al., 2021; Boulard et al., 2019). The resulting hypertrophic extracellular matrix may potentially contribute to the macroscopic muscle size, which would weaken the relation between macro- and microscopic muscle sizes. However, a hypertrophic extracellular matrix has been especially found in older children with CP with contractures, and is not yet confirmed in young ambulant children with CP. Our findings are in contrast to the results reported by Smith et al., 2011., who found smaller Fiber-CSA, but no change in diameter of the fascicles (i.e. bundles of muscle fibers) in the hamstrings muscle from children with CP compared to TD children. They explained this finding by the increase in epimysium collagen content in children with CP. Yet, the comparison between study results may not be valid, since the characteristics of the investigated children (especially their age) and the investigated muscles differed between this previous and the current study. For the current study sample of children with CP aged between 2 and 9 years, there are no data on the collagen content.

Next to the microscopic fiber size (Fiber-CSA), the within-patient variability in Fiber-CSA (CV) was significantly correlated with absolute values of MV and ML. However, this was only found for children with CP, who had higher CV values compared to TD children. Yet, these correlations were also low and the clinical meaning of these associations remains unclear. While the histological analyses on the same dataset revealed that children with CP had a significant higher percentage of smaller fibers (i.e. a fiber area between 0-750 µm2) compared to TD children (Deschrevel et al., 2023a), the averaged CSA for all fibers was not significantly different between both groups. Hence, it remains unclear which underlying microscopic structures may contribute to the macroscopic muscle size deficits in the investigated children with CP. One possible hypothesis for the reduced macroscopic muscle size in CP without a clear reduction in Fiber-CSA, could be that children with CP have fewer muscle fibers compared to TD children. A review on the mechanisms of muscle hypertrophy after training in healthy adults suggested that the increase in Belly-CSA in response to training results from fascicle growth due to fiber hypertrophy and/or due to an increase in the number of fibers per cross section, next to longitudinal fascicle growth (Jorgenson et al., 2020). Previous literature is characterised by inconsistent findings on the alterations of fascicle length in children with CP. Fascicle lengths were found to be similar or shorter in CP compared to TD muscles (Barber et al., 2011; Matthiasdottir et al., 2014; Mathewson et al., 2015; D’Souza et al., 2019;Handsfield et al., 2022). It is therefore plausible that children with CP have a fewer amount of muscle fibers and/or reduced capacity to regenerate muscle fibers. These microscopic alterations could potentially be ascribed to alterations in satellite cell amount or function, since satellite cells are responsible for postnatal growth and muscle regeneration. However, although reduced amounts of satellite cells have been found in different lower limb muscles other than the MG, especially in older children with muscle contractures (Smith et al., 2013; Dayanidhi et al., 2015; Kahn et al., 2023), no reductions in the number of satellite cells were observed in the MG of the younger ambulant children from the current study (Corvelyn et al., 2020; Deschrevel et al., 2023a). In general, it is important to acknowledge that only few studies have investigated the microscopic muscle size properties and related structures (such as the satellite cells and components of the extracellular matrix) of the MG in young children with CP. Moreover, it has been suggested that muscle growth is characterized by an uneven growth distribution (Jorgenson et al., 2020), highlighting the importance of the biopsy site.

In pennated muscles, such as the MG, the alterations in the macroscopic physiological Belly-CSA (i.e. muscle belly cross-sectional area perpendicular to the muscle fascicle direction), may be more directly related to the microscopic Fiber-CSA. Unfortunately, the current macroscopic dataset does not include the required architectural muscle parameters to estimate the physiological Belly-CSA (i.e. the pennation angle and fascicle length). However, previous studies showed no differences in the MG pennation angle between children with CP and TD children (D’Souza et al., 2019; Barber et al., 2011), and, as mentioned above, inconsistent findings have been found on the alterations of fascicle lengths. Combined with the observed low correlations between deficits in muscle volume (zMV) and deficits in the microscopic Fiber-CSA (zFiber-CSA), we consider it unlikely that alterations in the macroscopic physiological Belly-CSA would set more light on the complex interaction between the macro- and microscopic muscular structures.

We also explored possible correlations between macroscopic muscle parameters, including EI, and other microscopic muscle parameters, to further improve our understanding of the coherence between both. Concerning capillarisation, only capillary density was correlated with absolute values of ML in TD children. The negative correlation could point towards a less denser network along the muscle length in larger muscles, as capillary density decreases with increasing age. Additionally, all absolute macroscopic muscle size parameters (MV, Belly-CSA, ML) showed low to moderate significant correlations with the capillary to fiber ratio (C/F), both in children with CP as well as TD children. This finding may be explained by the expected increase in capillaries per fiber with increasing muscle size (Wüst et al., 2009; Ahmed et al., 1997; Bosutti et al., 2015). Indeed, when excluding the impact of anthropometric growth through the use of deficits (z-scores), the capillary density and C/F were not anymore significantly correlated with deficits (z-scores) of the macroscopic muscle size parameters. The fiber type proportion showed few and low to negligible correlations with absolute macroscopic muscle parameters.

To the best of our knowledge, this is the first study that explored the relation between macro- and microscopic muscle properties of the same muscle in one cohort of children with CP and TD children. The current findings highlight that knowledge on the microscopic muscular level cannot be simply transferred to the macroscopic level, and vice versa. A reduced macroscopic muscle size, such as a small muscle belly CSA or volume, does not necessarily indicate a reduced Fiber-CSA and thus also not a reduced number of sarcomeres in parallel. This may to some extent explain why muscle size only partly predict muscle strength and function in children with CP (Reid et al., 2015; Hanssen et al., 2021).

Some study limitations have to be acknowledged. First, because only children up to 9 years of age were included, our results cannot be generalized to older children, especially since puberty is characterized with many hormonal changes that may affect muscular growth (Gillen et al., 2021). For children with CP, solely ambulant children participated in this study. Hence, there is a need for further research in older and more affected children. Secondly, due to the large variability in fiber CSA within children with CP, the averaged fiber-CSA may not equally represent their microscopic muscle size compared to TD children. Therefore, we also defined the CV of the fiber CSA as a primary outcome. Thirdly, the existing literature on the underlying structures responsible for intrinsic muscular alterations is inconclusive, making it difficult to find explanations responsible for the limited interactions between macro- and microscopic muscular deficits. Finally, the current study primarily focused on muscle morphology. The knowledge on the intrinsic muscle composition, i.e. the relative content of contractile and non-contractile elements, is still scarce. It is generally accepted that muscle composition is altered in CP, due to increased fat infiltration and collagen content (Howard and Herzog, 2021). However, there is a lack of CP studies that provide quantitative information on muscle composition. More extensive research is thus needed to provide firm conclusions.

## 5. CONCLUSION

In conclusion, this integration of macro- and microscopic muscle (size) parameters in children with CP highlighted the complexity of the interacting network of macro- and microscopic muscle structures. While the absolute values for macro- and microscopic muscle size parameters were moderately correlated, their deficits (z-scores) relative to TD children revealed only one significant low correlation between zMV and zFiber-CSA. The increased inter-subject variability in Fiber-CSA (CV), which was clearly observed in children with CP compared to TD children, was only weakly correlated with the absolute macroscopic MV and ML parameters. Hence, it remains unclear how alterations in microscopic muscular structures contribute to the macroscopic muscle size deficits. Future studies covering a more extended set of macro- and microscopic muscle structures are recommended to unravel this complex interaction between macro- and microscopic muscle parameters in children with CP.

## Acknowledgments

We would like to thank all the children and their parents for participating in this study, as well as the colleagues of University Hospital Leuven for helping in the recruitment of children with CP and contributing to the biopsies. We thank all the students of the Clinical Motion Analysis Laboratory and the laboratory of Respiratory Diseases and Thoracic Surgery of the University of Leuven for the help in recruitment of the TD children and data collection.

This research was funded by an Internal KU Leuven grant 3D-MMAP (C24/18/103), and by funding from the Research Foundation Flanders (FWO) for the FWO research projects G0B4619N and G084523N, and the FWO research fellowship to co-author IV (1188923N).

## Author contribution

Conceptualization of this study was done by CL and KD. JD, AA, NDB, BH and IV participated in the data collection and processing. Data analysis and visualisation was performed by CL, supervised by KD. Data interpretation was done by CL, GGR, AVC and KD. Funding acquisition was acquired via GGR, AVC and KD. The manuscript was written by CL, while JD, AA, KM, NDB, BH, IV, AVC, GGR and KD reviewed and edited the manuscript. All authors have read and agreed to the published version of the manuscript

## Data availability statement

The datasets generated for this study can be found in the Research Data Repository (RDR) of the KU Leuven at …. (doi-link will be uploaded when paper is accepted)

