## Supplementary material for "The relation between macro- and microscopic intrinsic muscular alterations of the medial gastrocnemius in children with spastic cerebral palsy": Table S1

TABLE S1	Summary of the means and standard deviations of macroscopic parameters, and independent t-test outcomes

|  | TD | CP | | Independent t-test |
| --- | --- | --- | --- | --- |
|  | N mean ± SD | | N mean ± SD | t-value p-value |
| Absolute parameters |  | |  |  |
| MV (ml) | 31 46.0 ± 15.6 | | 42 25.7 ± 10.7 | -6.3 **p<0.001** |
| Belly-CSA (mm^2^) | 31 464.4 ± 103.3 | | 40 305.7 ± 93.1 | -6.8 **p<0.001** |
| ML (mm) | 33 156.4 ± 19.5 | | 42 126.3 ± 21.2 | -6.3 **p<0.001** |
| EI | 31 150.3 ± 11.9 | | 41 169.4 ± 14.2 | 6.1 **p<0.001** |
| Normalized parameters |  | |  |  |
| nMV (ml/kg*m) | 31 1.8 ± 0.23 | | 42 1.3 ± 0.33 | -7.5 **p<0.001** |
| nBelly-CSA (mm^2^/fibula^2^) | 31 0.74 ± 0.13 | | 40 0.60 ± 0.18 | -3.6 **p<0.001** |
| nML (mm/m) | 33 130.8 ±8 8.5 | | 42 116.4 ± 10.6 | -6.3 **p<0.001** |
| Z-scores (deficits) |  | |  |  |
| zMV |  | | 42 -2.10 ± 1.13 |  |
| zBelly-CSA |  | | 40 -2.08 ± 1.60 |  |
| zML |  | | 42 -1.18 ± 0.92 |  |

All macroscopic parameters are displayed as mean ± SD (standard deviation)

Abbreviations: TD, typically developing; CP, cerebral palsy; SD, standard deviation; MV, muscle volume; ml, millilitres; CSAb, cross-sectional area muscle belly; mm^2^, squared millimetre; ML, muscle length; mm, millimetre; EI, echo-intensity; nMV, normalized muscle volume; kg, kilogram; m, metre; nCSAb, normalized cross-sectional area of the muscle belly; nML, normalized muscle length; zMV, z-scores of muscle volume; zBelly-CSA, z-scores of muscle belly; zML, z-scores of muscle length

The critical p-value after Bonferroni correction is 0.05/7=0.007. Significant p-values are printed in bold.
