## Supplementary material for "The relation between macro- and microscopic intrinsic muscular alterations of the medial gastrocnemius in children with spastic cerebral palsy": Figure S1

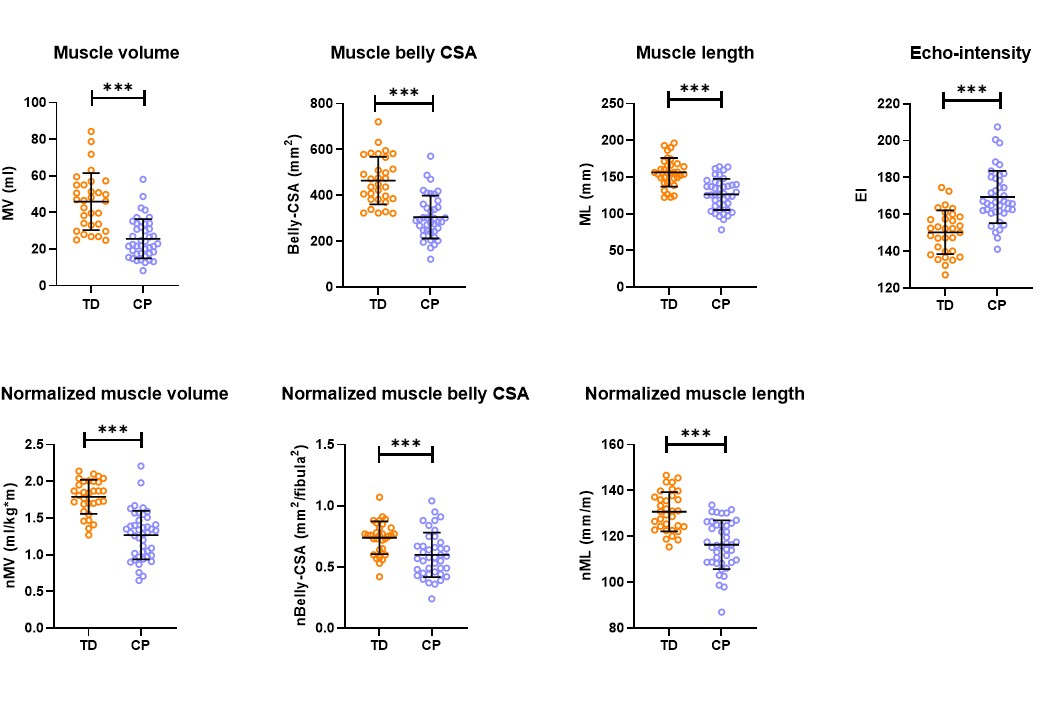

FIGURE S1 Individual datapoints for TD (orange) and CP (purple), per macroscopic parameter. Mean and standard deviation are given in black for each parameter. Group differences are displayed as *** (p<0.007).

Abbreviations: TD, typically developing; CP, cerebral palsy; MV, muscle volume; ml, millilitres; Belly-CSA, cross-sectional area of the muscle belly; mm², squared millimetres; ML, muscle length; mm, millimetres; EI, echo-intensity; nMV, normalized muscle volume; kg, kilogram; m, metres; nBelly-CSA, normalized cross-sectional area of the muscle belly; nML, normalized muscle length
