## Supplementary material for "The relation between macro- and microscopic intrinsic muscular alterations of the medial gastrocnemius in children with spastic cerebral palsy": Table S2

TABLE S2	Summary of the medians and interquartile ranges of the microscopic parameters, and Mann-Whitney U test outcomes

All microscopic parameters are displayed as median (interquartile range), except for z-scores (mean; standard deviation (SD))

Abbreviations: TD, typically developing; CP, cerebral palsy; MWU, Mann-Whitney U; IQR, interquartile range; Fiber-CSA, averaged cross-sectional area of all fibers; µm², squared micrometre; CV, coefficient of variation; C/F, capillary to fiber ratio; nFiber-CSA, normalized value of the cross-sectional area of all fibers; zFiber-CSA, z-score of the averaged cross-sectional area of all fibers; zCV, z-score of coefficient of variation; zCapillary density, z-score of capillary density; zC/F, z-score of capillary to fiber ratio

The critical p-value after Bonferroni correction is 0.05/7=0.007. Significant p-values are printed in bold.

|  | TD | CP | | MWU test |
| --- | --- | --- | --- | --- |
|  | N median (IQR) | | N median (IQR) | U-value p-value |
| Absolute parameters |  | |  |  |
| Fiber-CSA (µm^2^) | 34 1596.2 (737.9) | | 45 1106 (946.5) | 993.5 p=0.024 |
| CV (%) | 34 26.9 (7.8) | | 43 38.3 (11.1) | 177 **p<0.001** |
| Fiber type I proportion (%) | 34 66.5 (12.6) | | 45 51 (14.9) | 1210 **p<0.001** |
| Fiber type IIa proportion (%) | 34 27.7 (6.9) | | 45 25.1 (8.8) | 937 p=0.089 |
| Fiber type IIx proportion (%) | 34 5.6 (8.7) | | 45 22.8 (11.2) | 136 **p<0.001** |
| Capillary density (capillaries/mm^2^) | 27 493.9 (99.8) | | 38 365.9 (135.2) | 859 **p<0.001** |
| C/F | 27 1.4 (0.59) | | 38 0.76 (0.59) | 856 **p<0.001** |
| Normalized parameters |  | |  |  |
| nFiber-CSA (µm^2^/fibula^2^) | 34 2.3 (0.96) | | 43 2.3 (1.4) | 704 p=0.782 |
| Z-scores (deficits) |  | | N mean ± SD |  |
| zFiber-CSA |  | | 45 -0.14 ± 1.61 |  |
| zCV |  | | 43 1.85 ± 1.30 |  |
| zCapillary density |  | | 38 -2.26 ± 1.55 |  |
| zC/F |  | | 38 -2.08 ± 2.31 |  |
