## Supplementary material for "The relation between macro- and microscopic intrinsic muscular alterations of the medial gastrocnemius in children with spastic cerebral palsy": Figure S2

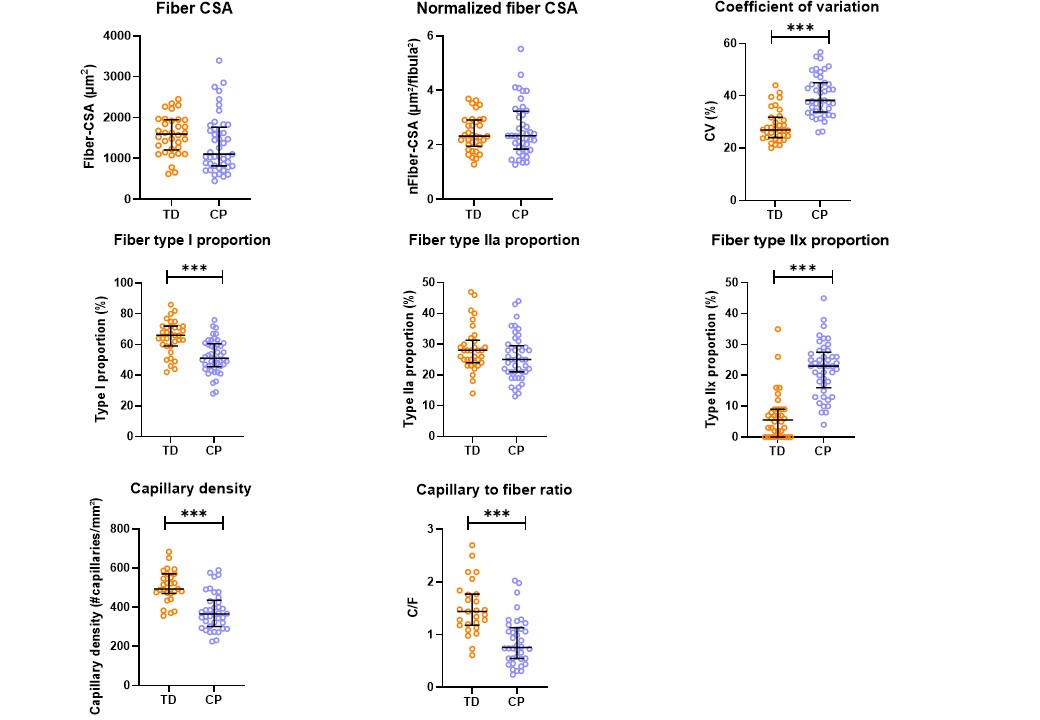

FIGURE S2 Individual datapoints for TD (orange) and CP (purple), per microscopic parameter. Median and interquartile range are given for each parameter. Group differences are displayed as *** (p<0.007).

Abbreviations: TD, typically developing; CP, cerebral palsy; Fiber-CSA, averaged cross-sectional area of all fibers; µm², squared micrometre; nFiber-CSA, normalized value of the cross-sectional area of all fibers; CV, coefficient of variation; C/F, capillary to fiber ratio
